# Patient and Practice Level Visual Acuity Prior to Cataract Surgery: An IRIS^®^ Registry (Intelligent Research in Sight) Analysis

**DOI:** 10.1101/2025.07.07.25331037

**Authors:** Laurel Tainsh, Vivian Paraskevi Douglas, Joshua B. Gilbert, Sarah Manz, Connor J. Ross, William Kearney, Tobias Elze, Joan Miller, Alice Lorch

## Abstract

**Purpose:** To examine the influence of patient demographic characteristics and ophthalmic practice composition on access to cataract surgery in the United States as measured by preoperative best-corrected visual acuity (BCVA).

**Patient and methods:** This retrospective cohort study analyzed data from the IRIS^®^ Registry (Intelligent Research in Sight) for patients age > 50 who had at least one BCVA measurement in the six months preceding cataract surgery performed between January 1, 2016, and December 31, 2020. We used a mixed-effects model to estimate the relationship between individual-level demographic factors and practice-level composition factors and preoperative BCVA.

**Results:** 2,387,045 individuals met inclusion criteria. The mean BCVA prior to surgery was 0.23 (SD: 0.32) logMAR. The worst pre-operative BCVA was observed in patients with Hispanic race and ethnicity while White patients had the best [0.34 (SD: 0.43), 0.21(SD: 0.30); p<0.001]. Grouping patients in terms of percentage of BCVA worse than 20/50 prior to surgery, Hispanic patients, active smokers, and uninsured patients had higher percentages of worse preoperative vision (33.7%, 23.5%, 34.9%). Analysis of compositional effects of race and ethnicity, smoking and insurance status showed that, regardless of an individual patient’s demographic, patients treated at practices serving higher proportions of White patients showed better BCVA (b = −.008 per 10 percentage points, *P* < .001) while patients at practices with higher percentages of actively smoking patients showed worse BCVA (b=-0.016 per 10 percentage points active smoking patients, *P* < .001). There was no compositional effect of insurance status.

**Conclusions and Relevance:** Overall differences exist with regard to the visual acuity at which cataract surgery is initiated at both the level of the individual patient and the composition of practice in which they are treated.

**Plain Language Summary:** Demographic disparities and geographic variation in access to cataract surgery in the United States have been previously described in large national studies of insurance data. Smaller studies of single institutions expanded upon these studies by showing differences in preoperative visual acuity- an important measure of access to cataract surgery- based on factors such as race and insurance status but were limited by the size and scope of their study patients. The IRIS^®^ Registry (Intelligent Research in Sight) is the nation’s first comprehensive ophthalmic clinical registry with data from both individual patients as well as ophthalmic group practices. Using data from this registry, we show differences in preoperative visual acuity prior to cataract surgery at both the level of the patient and the practice in which they are treated.

## Introduction

Cataract is the leading cause of visual impairment worldwide and cataract extraction is the most commonly performed procedure in the United States with increasing rates over the past few decades.^1–3^ As cataract becomes more prevalent and surgery becomes safer and more efficient, the visual threshold to operate has declined to accommodate the aging population’s increased visual demands.^4^ Access to cataract surgery, however, has been historically inequitable with many prior studies demonstrating disparities in access to and timing of cataract surgery based on demographic factors.^5–9^

The American Academy of Ophthalmology IRIS^®^ Registry (Intelligent Research In Sight) database provides an important opportunity to explore disparities in access to cataract surgery as measured by best corrected visual acuity (BCVA) prior to surgical intervention. While previous studies demonstrated worse preoperative visual acuity (VA) in patients with varied race and insurance status, these studies were typically institution or region based and therefore limited in the size and scope of cohort.^8^ Larger national studies exist, but have relied on insurance claims that lack clinical data such as VA instead relying on indirect metrics such as patient age at surgery and time from ICD code diagnosis to surgery.^6,9^ Prior studies also typically model data at the level of the patient and ignore the important contextual effect of practice demographic composition.^4,7^ In this study, we use the IRIS Registry to explore demographic factors associated with BCVA prior to cataract surgery nationwide at both the level of the individual patient and the demographic composition of the practice in which they are treated and expand our understanding of access to cataract surgery in the United States.

## Methods

We used a retrospective cohort design with de-identified electronic health record (EHR) data of patients followed at practices that participate in the IRIS Registry. The version of the database was frozen on December 24, 2021 and accessed on July 9, 2023. The data collection methodology of the IRIS Registry has been described previously.^10^ The investigation was approved by the Massachusetts General Brigham Institutional Review Board with the exemption of informed consent due to the anonymized and retrospective nature of the analysis. The research adhered to the Declaration of Helsinki.

We included the records for patients aged over 50 who underwent cataract surgery between January 1, 2016 and December 31, 2020, and had at least one VA measurement from the eye on which cataract surgery was performed within the six months prior to surgery. Cataract surgery was identified using Current Procedural Terminology (CPT) codes 66982, 66983, and 66984. Any cataract surgery missing a CPT code specification for laterality was excluded in order to match the BCVA observations to the laterality of the eye on which the surgery was performed. Only the first cataract surgery on the first eye recorded in the dataset was included because preoperative BCVA could be affected by a history of prior cataract surgery. Patients with diagnoses that could affect vision independent of cataract were excluded including diabetic retinopathy (ICD-10 codes E10.3X, E11.3X), diagnoses involving the retina (ICD-10 codes H31.X, H33.X, H34.X, H35.X, H43.X, H44.X), diagnoses involving the optic nerve (ICD-10 codes H46.X, H47.X), and diagnosis of sudden vision loss (ICD-10 codes H53.X), with *X* representing any number of alphanumeric characters. We determined race and ethnicity based on EHR documentation. We excluded all persons without race or ethnicity information, except for patients identified as Hispanic ethnicity but lacking other race and ethnicity information.

### Outcome Measures and Variables

The outcome was the mean BCVA of the eye undergoing surgery at visits within the six months before surgery to approximate the eye’s preoperative BCVA. BCVA was defined as the lowest (“best”) logarithm of the minimum angle of resolution (logMAR) value available among all the viable observations for that eye, including those with or without pinhole VA, with or without old or new refraction, and at any distance.^11^ LogMAR values measured with Brightness Acuity Testing were not used. Visual acuity was extracted in logMAR, rather than Snellen, for the purposes of statistical analysis. A 0.1 change in logMAR is equal to a one-line change on the Snellen eye chart.

Race and ethnicity were operationalized as the following set of mutually exclusive categories: Asian, Black or African American (hereafter, Black), Hispanic, Non-Hispanic White (hereafter, White), and Other (including Multiracial, Native American/Alaska Native, Native Hawaiian/Pacific Islander). Patients were assigned the above categories such that race (Asian, Black, Other) superseded ethnicity (Hispanic), except for White race. As such, a patient with Black race and Hispanic ethnicity would be categorized as Black.

### Statistical Analysis

To estimate the relationship between race and ethnicity and preoperative BCVA, both within and between ophthalmology practices, we used a series of single- and multi-level (mixed effects) regression models. Model 1 used a simple linear regression model of BCVA on race and ethnicity without additional covariates to determine average differences across racial groups as a baseline for comparison. Model 2 added a random effect for ophthalmology practice to account for clustering of patients within the same practice. Model 3 added practice-level means of patient covariates (race and ethnicity, smoking status, and insurance status) as well as practice size to account for systematic differences between practices and provide the contextual effects of demographic composition. The contextual variables were multiplied by 10 so that the coefficient provides the difference in BCVA per 10 percentage point difference in the contextual variable. Model 4 added patient-level covariates to adjust for additional individual-level differences between racial and ethnic groups thus reducing the effects of confounding variables.

The patient-level coefficients of interest were the racial and ethnic group variables, which reflect the within-practice differences in BCVA between patients identified as Asian and patients in all other racial and ethnic groups, holding the other covariates constant. Asian race and ethnicity were used as the basis of comparison by alphabetical convention per previously described guidelines.^12^ The practice-level coefficients of interest were the racial and ethnic composition variables, which represent the between-practice (contextual) differences in BCVA predicted by the differing racial and ethnic compositions of each practice’s patient population, holding individual race and ethnicity constant. In other words, the within-practice effects compare the BCVA of patients of different racial and ethnic groups served by the same practice, whereas the between-practice effects compare the average BCVA of practices with different racial and ethnic compositions, adjusting for the other variables in the model. Reference groups for each covariate are described in **Table 3**.

All statistical tests were two-sided, and we set the threshold for statistical significance at *P* < 0.05. All analyses were performed using R version 4.4.2.

## Results

A total of 2,387,045 individuals met inclusion criteria for the study (**Table 1**). The mean age of the population was 70.6 [standard deviation (SD) = 8.2]. A majority of patients identified as female (59%), having private insurance (59%), and as never smokers (54%). A large majority of patients were White (83.0%). The mean BCVA prior to surgery in logMAR was 0.23 SD: 0.32 (approximately 20/32 Snellen) and the average time of recorded BCVA before surgery was 1.38 (SD: 1.18) months (**Table 1**).

**Table 1.** Demographic and clinical characteristics of the cohort.

|  |  |
| --- | --- |
| Characteristic | **N = 2,387,045** |
| Age (decades over 50 years old) | 2.06 (0.82) |
| Sex |  |
| **Female** | 1,405,904 (59%) |
| **Male** | 981,141 (41%) |
| **Medicaid patient\*** | 217,883 (9.1%) |
| *Insurance status* |  |
| **Government** | 51,497 (2.2%) |
| **Medicare** | 670,745 (28%) |
| **Military** | 87,323 (3.7%) |
| **Private** | 1,403,479 (59%) |
| **Uninsured** | 3,363 (0.1%) |
| **Unknown** | 170,638 (7.1%) |
| *Smoking status* |  |
| **Active** | 277,675 (12%) |
| **Former smoker** | 587,966 (25%) |
| **Never** | 1,279,477 (54%) |
| **Unknown** | 241,927 (10%) |
| *Region* |  |
| **North Central** | 557,381 (23%) |
| **Northeast** | 271,211 (11%) |
| **South** | 1,058,084 (44%) |
| **West** | 500,369 (21%) |
| *Race/Ethnicity* |  |
| **Asian** | 57,184 (2.4%) |
| **Black** | 202,122 (8.5%) |
| **Hispanic** | 115,679 (4.8%) |
| **Other** | 41,687 (1.7%) |
| **White** | 1,970,373 (83%) |
| BCVA (logmar) | 0.23 (0.32) |
| Time pre-surgery (months) | 1.38 (1.18) |
| **Mean (SD); n (%)** |  |
| **\*in the version of the IRIS Registry used in our study data for Medicaid status and** |  |

**Table 2** shows the distribution of covariates by race and ethnicity. There were statistically significant differences across groups for all covariates due to large sample size (*P* < 0.001 for all). Age was clinically similar between the groups. There were more female than male patients in each racial and ethnic group. White patients had the highest percentage of private insurance (60%) and the least percentage of Medicaid (6.4%) while rates of other insurance statuses were similar across groups. The lowest percentage of active smokers was seen in Asian patients (5.5%) and highst in White and Black patients (12%). Hispanic patients had the highest pre-operative logMAR BCVA while White patients had the lowest [0.34 (SD: 0.43), 0.21(SD: 0.30); p<0.001] and therefore better vision (20/40 and 20/32 Snellen vision respectively). White patients had the shortest time between BCVA measurement and surgery while Hispanic patients had the longest [1.36 (SD: 1.16), 1.60 (1.32), p<0.001].

**Table 2.**
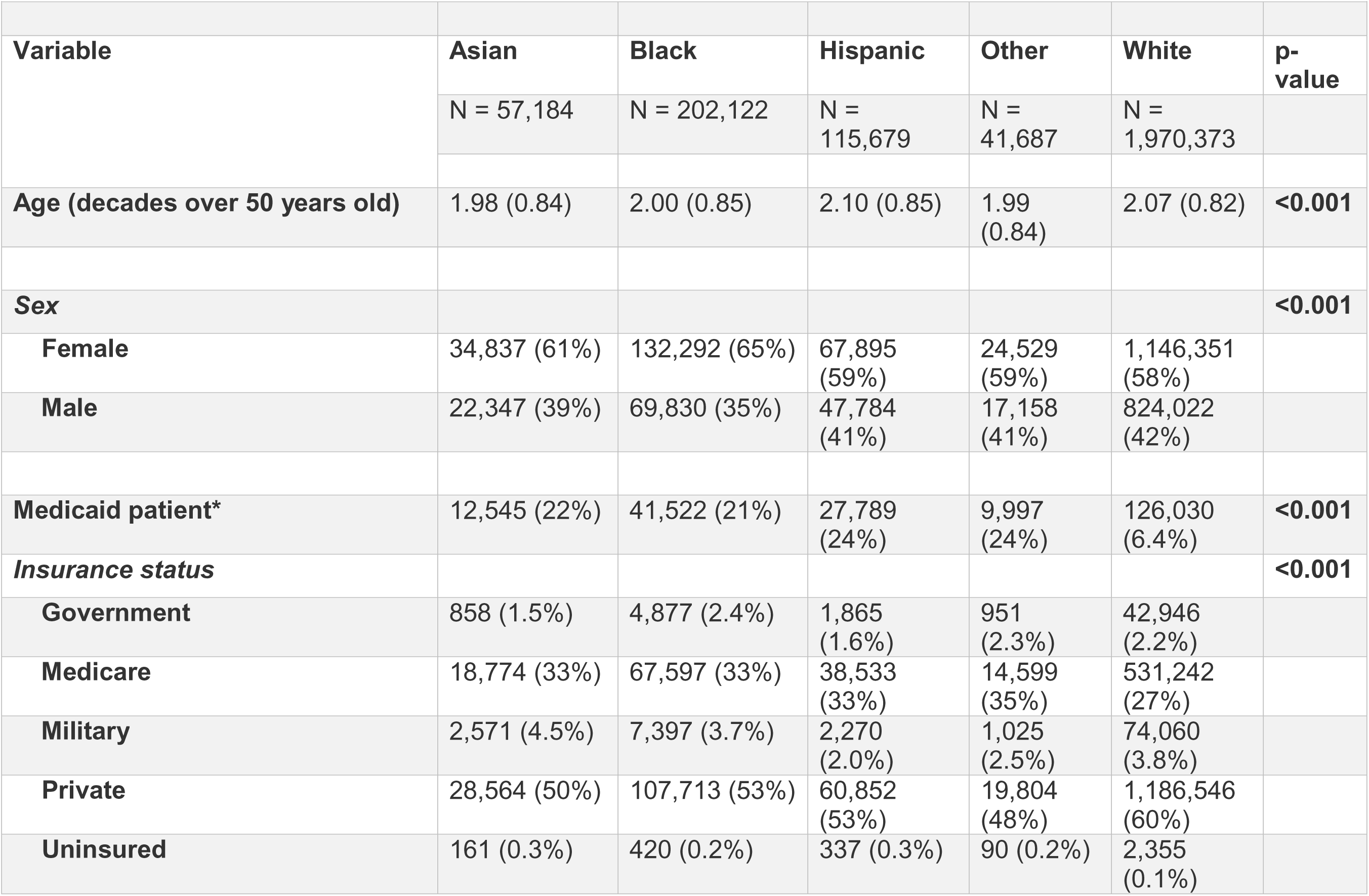
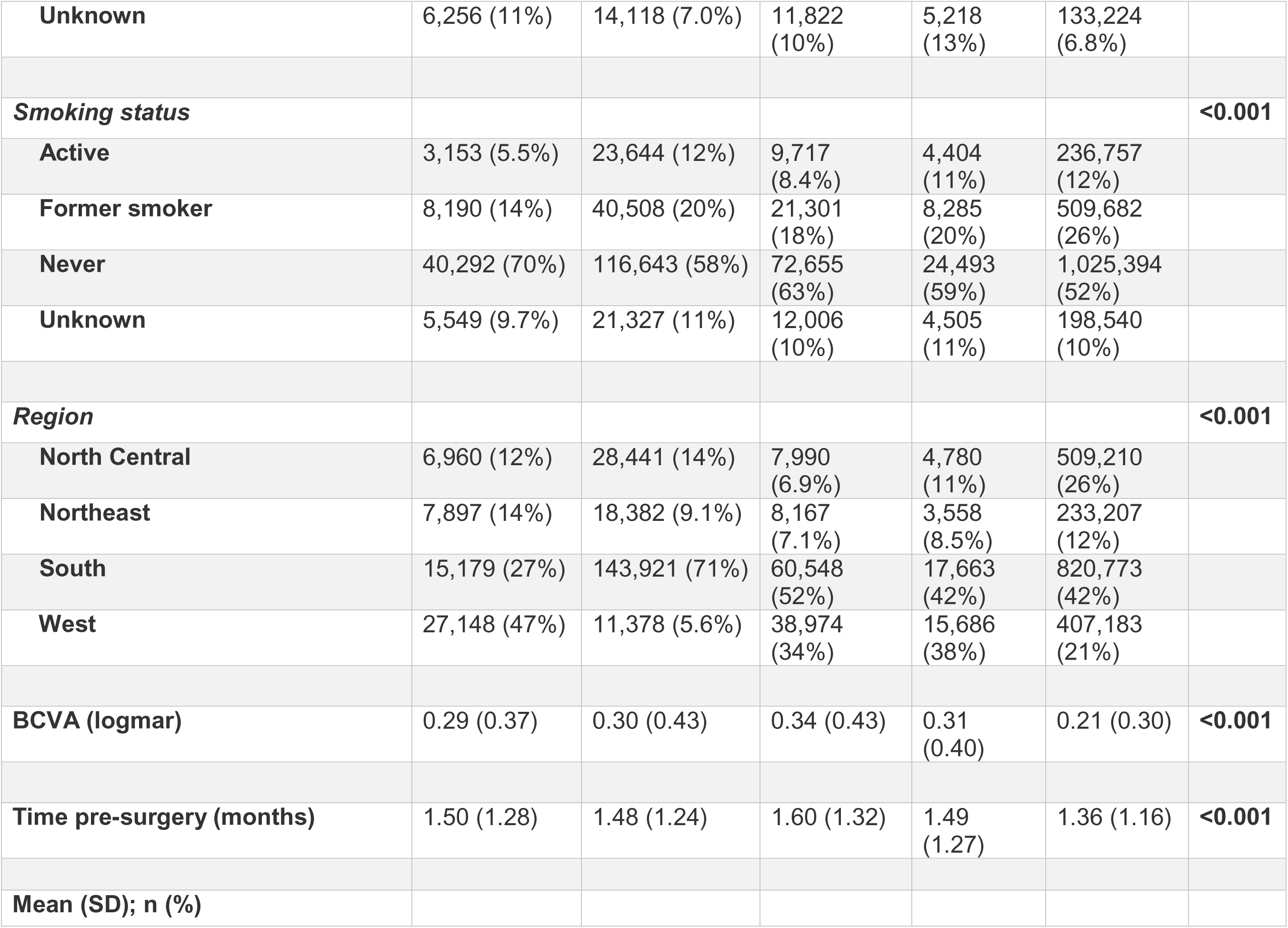
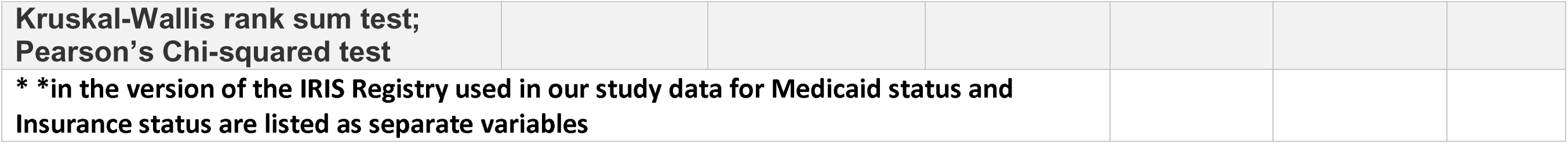
Distribution of covariates by race/ethnicity.

To more clearly illustrate clinically meaningful differences in pre-operative BCVA across covariates, we grouped patients in terms of percentages of patients with pre-operative vision better or worse than 20/50 for different race/ethnicity, insurance, and smoking statuses (**Figure 1**). Patients with Hispanic race/ethnicity had the highest percentage of pre-operative BCVA worse than 20/50 (33.7%) while White patients had the lowest (16.2%). Active smokers and uninsured patients also had higher percentages of BCVA worse than 20/50 prior to surgery (23.5%, 34.9%).

**Figure 1.**
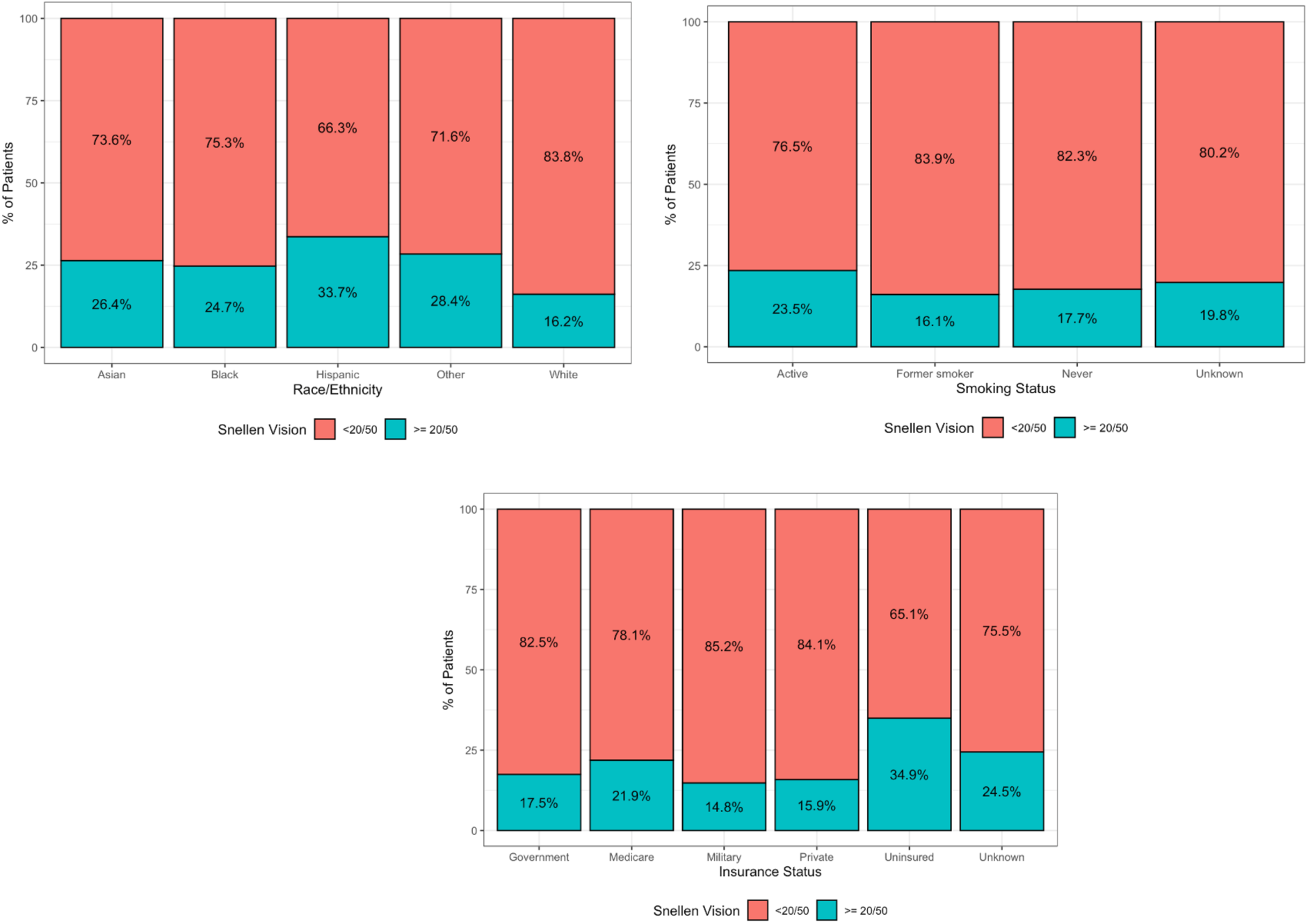
Proportion of patients in each best corrected visual acuity (BCVA) grouping by race/ethnicity, smoking status and insurance status

**Table 3** shows the results of the regression models. Model 1 shows small but statistically significant differences between racial and ethnic groups, with White patients reporting the best pre-operative BCVA (b= −0.078, p<.001 as compared to Asian patients, approximately 1 line of Snellen acuity). Model 2 adds the random effect for practice and provides an estimate of racial and ethnic group differences comparing individuals in the same practice. Model 2 shows substantial variation between practices: about 16% of the total variation in BCVA is at the practice level. We also see that the coefficients on racial and ethnic group are attenuated compared to Model 1 (e.g., White vs. Asian b = −.065), suggesting that some of the overall differences between groups observed in Model 1 are attributable to practice effects. Model 3 includes the practice-level variables and shows small but significant contextual effects (e.g., −.008 for each 10 percentage points of White patients vs. Asian patients, <1 line of Snellen acuity), and Model 4 includes the patient-level variables.

**Table 3.**
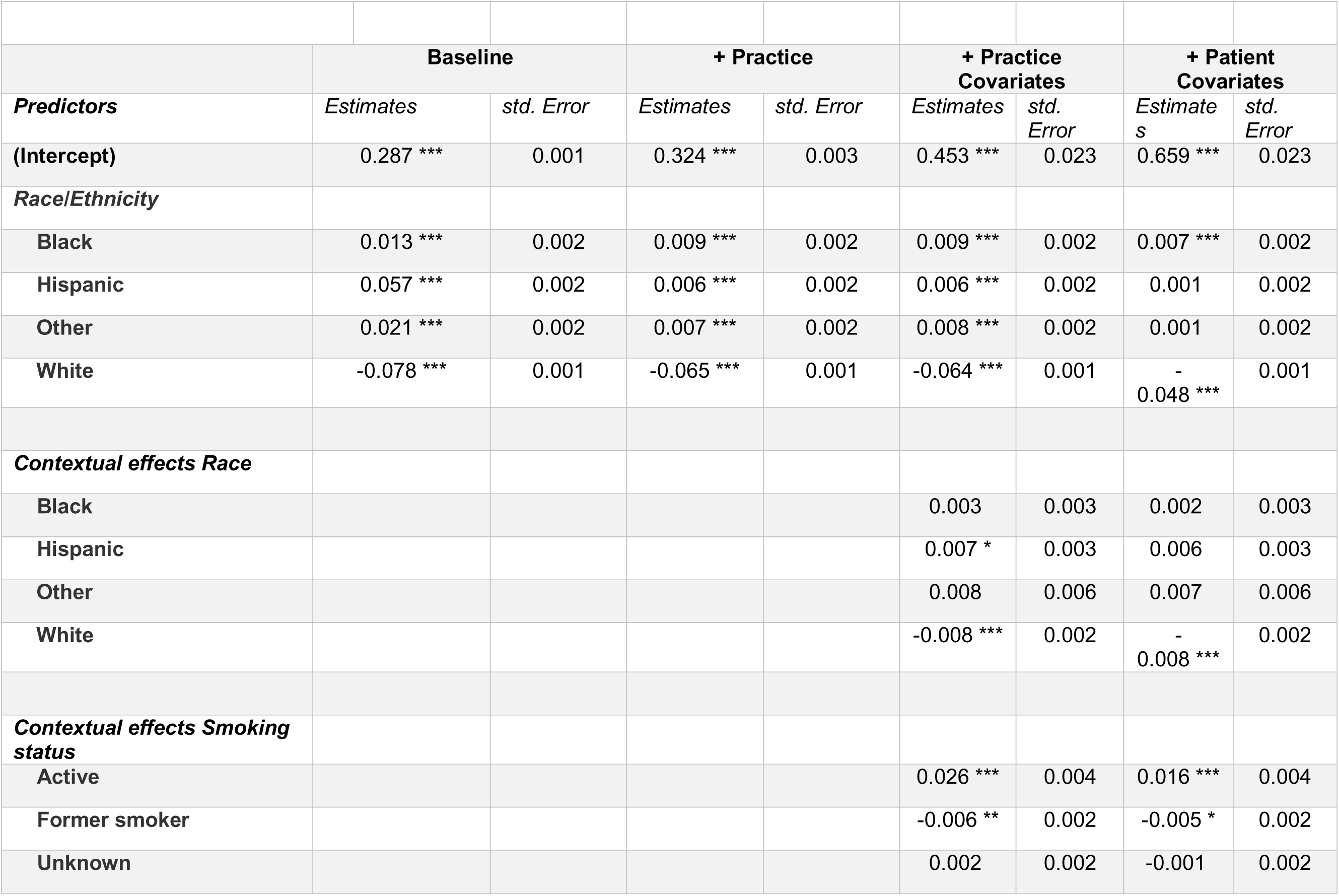
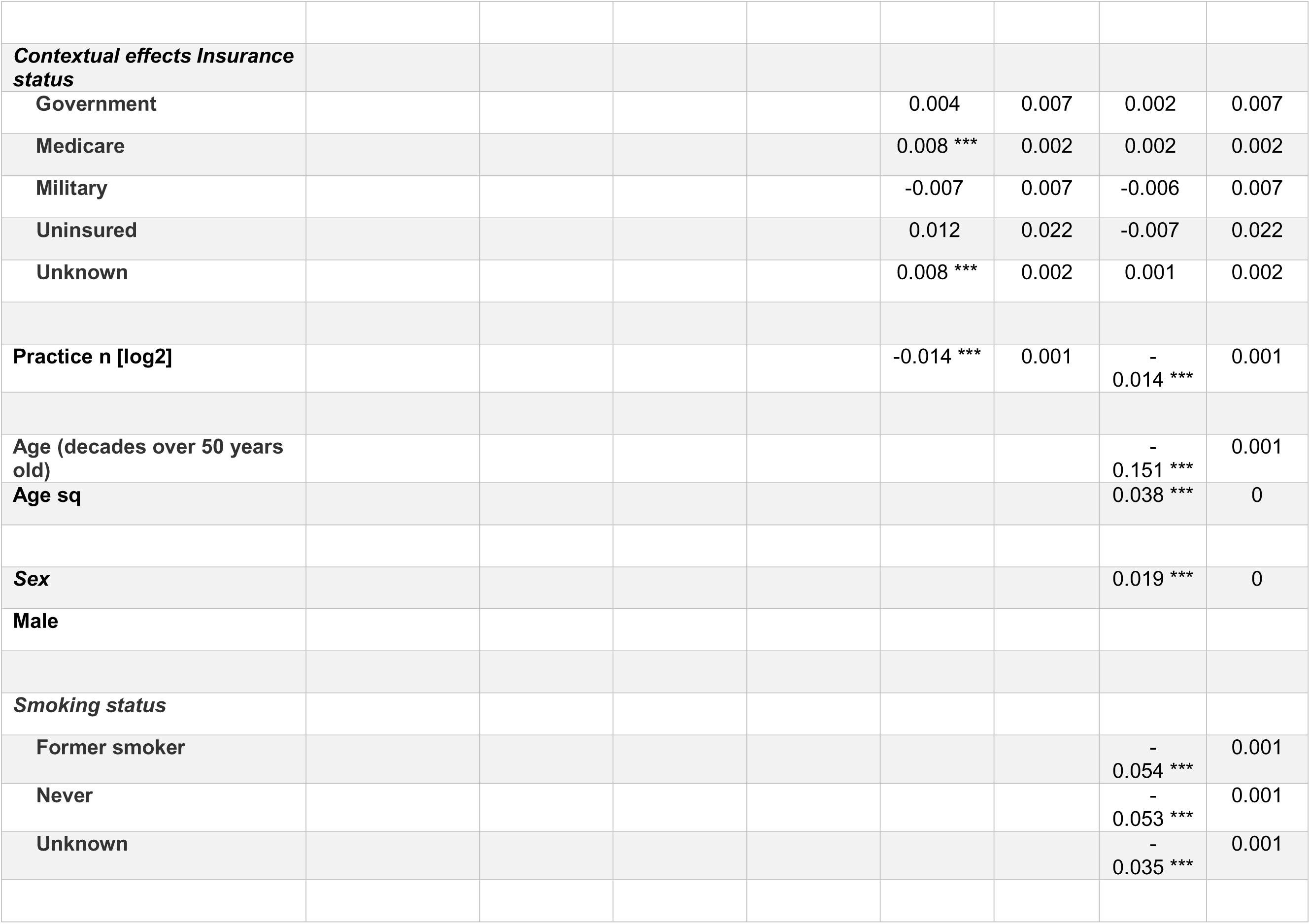
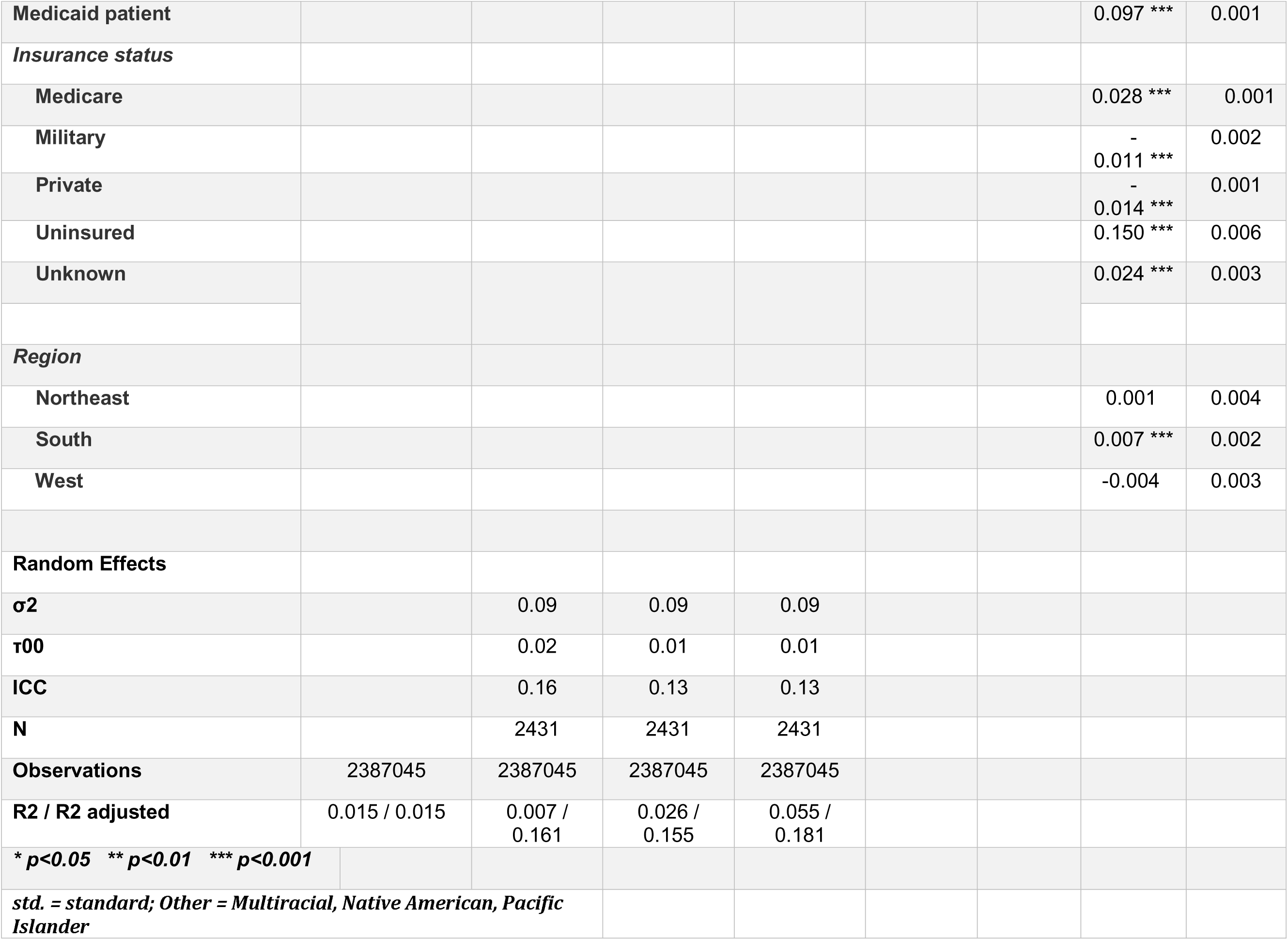
Linear mixed effects model: contribution of different demographic covariates to the BCVA of patients prior to cataract surgery, a within and between practice analysis.

The results of Model 4 provide the following insights. When controlling for practice and patient characteristics, the differences between Asian and Hispanic and Asian and Other race patients are near 0, in contrast to the baseline Model 1 that showed larger, significant differences, suggesting the importance of controlling for confounding variables. White patients continue to show better BCVA compared to Asian patients, but the adjusted difference is smaller in magnitude (b = −.048, *P* < .001, approximately ½ line of Snellen acuity). Interestingly, the contextual effect of practice composition for White patients is also negative and statistically significant (b = −.008, *P* < .001). This coefficient means that, if we take two patients of the same race and ethnicity, but the composition of their practices differs by 10 percentage points of White patients, the patient at the practice that serves more White patients reports better BCVA. Significant results in Model 4 were also found for the contextual effect of smoking status (practice level mean number of active smokers b=0.016 per 10 percentage points, *P* < .001), practice size (b= −0.014 per doubling of practice size, P < .001), and select patient level covariates for insurance status, smoking status, and geographic region (**Table 3**). The contextual effect of practice composition of insurance status was not significant in Model 4.

Figure 2 demonstrates the contextual effects of Model 4 in graphical format. As the proportion of Black, Hispanic, and Other race and ethnicity patients increases within a practice, the average pre-operative BCVA worsens but is not statistically significant.

**Figure 2.**
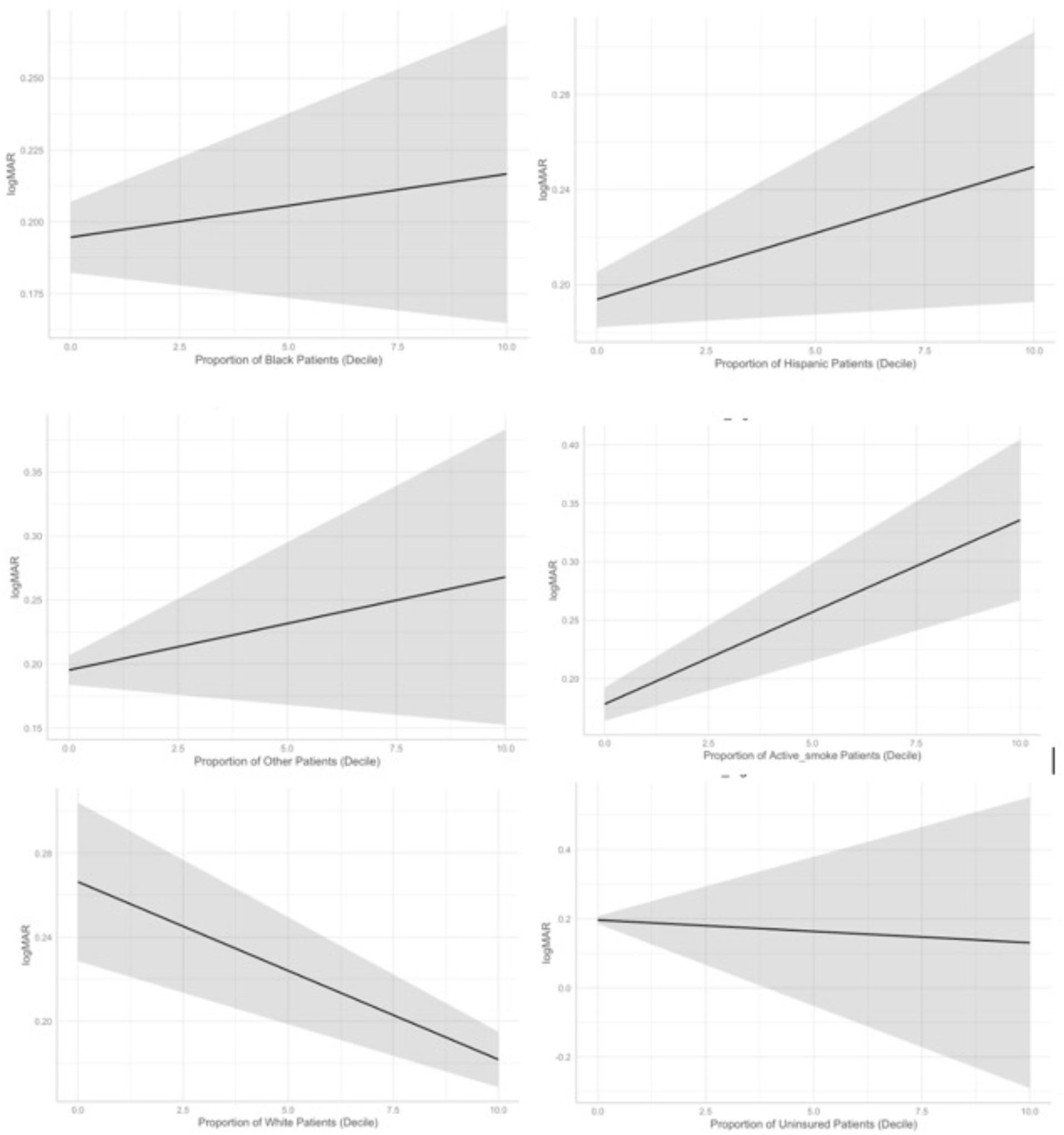
Proportion of patients in decile and predicted values of BCVA in logMAR

However, as the proportion of White patients in a practice increases, the preoperative BCVA improves in a small but significant manner. The proportion of actively smoking patients in a practice also corresponds to statistically significantly worse preoperative vision while the proportion of uninsured patients in a practice does not predict preoperative BCVA.

## Discussion

Our study was based on data from 2,387,045 patients aged 50 years and over in the IRIS Registry who underwent cataract surgery between 2016 and 2020, had at least one VA measurement within six months prior to surgery, and who were not excluded due to missing information or specific diagnoses that could affect vision potential independent of cataracts. We observed that the mean pre-cataract BCVA in our population was 0.23 logMAR (20/32 Snellen) taken at a visit on average 1.38 months prior to surgery.

Before controlling for the random effects of practice or for the contextual effects of practice composition, we observed better preoperative vision in White patients (Snellen 20/32 vs approx. 20/40-20/50 for other groups). Furthermore, we found a higher proportion of White patients with a preoperative BCVA of 20/50 or better and a lower proportion of BCVA 20/50 or better in actively smoking patients and uninsured patients. This finding has important practical implications because the minimum BCVA for obtaining or maintaining a driving license is 20/40 in almost all states nationally and a threshold of 20/40 vision has historically been used by insurance providers in determining eligibility for reimbursement for cataract surgery.^13,14^ Overall our findings suggest that patients who identify as White, with insurance, and who are not actively smoking receive surgical intervention at a point of better BCVA than their peers.

Previous studies also demonstrated disparities in ophthalmic care based on sociodemographic factors including race and ethnicity, and insurance status.^15–17^ Awidi et al showed worse preoperative BCVA in Black, Asian, and Hispanic patients prior to cataract surgery in two large US health systems.^18^ Similarly, a single-center study using data from 600 patients showed significantly worse pre-operative BCVA in patients of Hispanic race as well as in those patients with public insurance.^13^ The observations of our study expand upon these findings to a large national cohort.

To further understand observed potential disparities in access to cataract surgery, we used a mixed effects model to estimate the effects simultaneously of patient-level factors and the compositional effects of practice-level factors. Our results suggest that individual patients with race/ethnicity other than White or who are actively smoking were more likely to have worse pre-operative BCVA by about a ½ line of Snellen and uninsured and Medicaid patients by a full Snellen line (Table 3 Model 4 patient level covariates) compared to their respective reference groups. We also found that the average preoperative BCVA of a patient in a practice with a higher relative proportion of White patients and lower relative proportion of actively smoking patients were more likely to have better pre-operative BCVA though by a much smaller amount (<1 Snellen line for Table 3, model 4 contextual effects). There was no effect of practice composition in terms of insurance status, despite our findings for individual insurance status.

To our knowledge, our study is the first to examine the effect of practice demographic composition on access to cataract surgery. Using insurance claim data, Kauh et al examined the role of geographic variation in cataract surgery and found varying rates and timing of surgery in US cities and in rural vs urban practice location.^7^ While they did find varying hazards of cataract surgery in terms of race and ethnicity at the level of the individual, they did not consider the contextual effects of racial or other demographic composition in their model and did not look at the level of the individual practice ^7,13^. In contrast, a recent study in the urologic literature used a mixed effects model similar to our study to evaluate the role of practice organization and racial and ethnic composition in prostate cancer treatment and found that Black patients in practices with predominantly White patients underwent treatment at lower rates than Black patients in practices with predominantly Black patients.^19^ Similarly, in the cardiovascular surgery literature, Chen et al found that hospitals with a higher Racial/Ethnic Diversity Index experienced worse clinical outcomes and higher costs after aortic valve replacement.^20^ Our study adds to this important literature on the impact of clinic and hospital demographic composition and quality of care.

In sum, the statistical analyses show that while controlling for a host of variables reduces observed differences in BCVA between demographic groups, they are not eliminated. The significance of this effect suggests the presence of some sorting effects among practices and highlights the complexity underlying disparities in access to cataract surgery. For example, it may be that race and ethnicity, insurance, and smoking status are not the relevant factors, but an unobserved correlated variable such as income or education are the relevant factors. ^17^ The decision when to operate on a cataract is also complex and could depend on surgeon or practice specific decision making, visual demands of the population served by the practice, and individual patient motivation for surgery such as decline in visual function and effect on activities of daily living that are not measured by visual acuity alone such as glare and contrast sensitivity. ^9,21,22^ While we controlled for a rich set of variables, future studies should continue to explore the potential mechanisms and causes of these differences and the role of social determinants of health in access to cataract surgery.

There are several limitations of the data and study design to consider when interpreting our findings. Although our study sample represents a large cohort, certain geographic regions and racial and ethnic groups may be underrepresented in the IRIS Registry, thus limiting the generalizability of our results. The sex, race, and ethnicity groupings are based on what is documented in the EHRs and could be subject to reporting or inputting errors. Importantly, we acknowledge that race and ethnicity are cultural constructs and that the racial categories investigated in this study do not have well-defined nor clear scientific meanings. Estimates of a patient’s BCVA prior to cataract surgery were based on practice data in the IRIS Registry and could be subject to survivorship bias. There also may be variability in how VA was measured and considered across practices and in the quality of refraction. Nevertheless, the BCVA used in this study was likely representative of the measurement being used for surgical decision-making, given that it closely preceded surgery.

## Conclusions

In summary, rates of cataract surgery in the U.S. have increased in the past few decades, but disparities may exist with regard to the VA at which cataract surgery is initiated. These potential disparities exist at both the level of the individual patient and the patient population composition of the practice in which they are treated. Further work is needed to identify the causes behind this study’s findings and if similar patterns exist in ophthalmic practice beyond cataract surgery with the ultimate goal of reducing disparities in access to ophthalmic care.

## Data Availability

All data referenced in this manuscript are derived from the American Academy of Ophthalmology IRIS Registry. The Registry is a de‑identified, HIPAA‑compliant clinical database comprising tens of millions of patient encounters and accessible to researchers via AAO-approved pathways.

## References

1. Shu Y, Shao Y, Zhou Q, et al. Changing Trends in the Disease Burden of Cataract and Forecasted Trends in China and Globally from 1990 to 2030. Clin Epidemiol. 2023;15:525. doi:10.2147/CLEP.S404049

2. Chen X, Xu J, Chen X, Yao K. Cataract: Advances in surgery and whether surgery remains the only treatment in future. Advances in Ophthalmology Practice and Research. 2021;1(1):100008. doi:10.1016/J.AOPR.2021.100008

3. Erie JC. Rising cataract surgery rates: Demand and supply. Ophthalmology. 2014;121(1):2–4. doi:10.1016/j.ophtha.2013.10.002

4. Taylor HR, Vu HTV, Keeffe JE. Visual Acuity Thresholds for Cataract Surgery and the Changing Australian Population. Archives of Ophthalmology. 2006;124(12):1750–1753. doi:10.1001/ARCHOPHT.124.12.1750

5. Lange N, Kujawska-Danecka H, Wyszomirski A, et al. Significant improvements in cataract treatment and persistent inequalities in access to cataract surgery among older Poles from 2009 to 2019: results of the PolSenior and PolSenior2 surveys. Front Public Health. 2023;11:1201689. doi:10.3389/FPUBH.2023.1201689/BIBTEX

6. Schein OD, Cassard SD, Tielsch JM, Gower EW. Cataract surgery among Medicare beneficiaries. Ophthalmic Epidemiol. 2012;19(5):257–264. doi:10.3109/09286586.2012.698692

7. Kauh CY, Blachley TS, Lichter PR, Lee PP, Stein JD. Geographic Variation in the Rate and Timing of Cataract Surgery Among US Communities. JAMA Ophthalmol. 2016;134(3):267–276. doi:10.1001/JAMAOPHTHALMOL.2015.5322

8. Cho WKT, Hwang DG. Sociodemographic Disparities in Preoperative Visual Acuity and Cataract Surgery Utilization in the San Francisco Bay Area. J Racial Ethn Health Disparities. Published online February 8, 2024:1–14. doi:10.1007/S40615-024-01914-4/TABLES/5

9. French DD, Margo CE, Behrens JJ, Greenberg PB. Rates of Routine Cataract Surgery Among Medicare Beneficiaries. JAMA Ophthalmol. 2017;135(2):163–165. doi:10.1001/JAMAOPHTHALMOL.2016.5174

10. Chiang MF, Sommer A, Rich WL, Lum F, Parke DW. The 2016 American Academy of Ophthalmology IRIS ® Registry (Intelligent Research in Sight) Database: Characteristics and Methods. Ophthalmology. 2018;125(8):1143–1148. doi:10.1016/J.OPHTHA.2017.12.001

11. Brant A, Kolomeyer N, Goldberg JL, et al. Evaluating Visual Acuity in the American Academy of Ophthalmology IRIS® Registry. Ophthalmology Science. 2024;4(1):100352. doi:10.1016/J.XOPS.2023.100352

12. Flanagin A, Frey T, Christiansen SL. Updated Guidance on the Reporting of Race and Ethnicity in Medical and Science Journals. JAMA. 2021;326(7):621–627. doi:10.1001/JAMA.2021.13304

13. Stone JS, Fukuoka H, Weinreb RN, Afshari NA. Relationship Between Race, Insurance Coverage, and Visual Acuity at the Time of Cataract Surgery. Eye Contact Lens. 2018;44(6):393–398. doi:10.1097/ICL.0000000000000443

14. Legal Vision Requirements for Drivers in the United States. AMA J Ethics. 2010;12(12):938–940. doi:10.1001/VIRTUALMENTOR.2010.12.12.HLAW1-1012

15. Broman AT, Hafiz G, Muñoz B, et al. Cataract and barriers to cataract surgery in a US Hispanic population: Proyecto VER. Arch Ophthalmol. 2005;123(9):1231–1236. doi:10.1001/ARCHOPHT.123.9.1231

16. Richter GM, Chung J, Azen SP, Varma R. Prevalence of visually significant cataract and factors associated with unmet need for cataract surgery: Los Angeles Latino Eye Study. Ophthalmology. 2009;116(12):2327–2335. doi:10.1016/J.OPHTHA.2009.05.040

17. Morales LS, Varma R, Paz SH, et al. Self-reported use of eye care among Latinos: the Los Angeles Latino Eye Study. Ophthalmology. 2010;117(2). doi:10.1016/J.OPHTHA.2009.07.015

18. Awidi AA, Woreta FA, Sabit A, et al. The Effect of Racial, Ethnic, and Socioeconomic Differences on Visual Impairment before Cataract Surgery. Ophthalmology. 2025;132(1). doi:10.1016/J.OPHTHA.2024.07.021

19. Agochukwu-Mmonu N, Qin Y, Kaufman S, et al. Understanding the Role of Urology Practice Organization and Racial Composition in Prostate Cancer Treatment Disparities. JCO Oncol Pract. 2023;19(5):e763–e772. doi:10.1200/OP.22.00147

20. Chen Y, Xiao Y, Huang R, et al. Association between hospital racial composition and aortic valve replacement outcomes: A national inpatients sample database analysis. Catheter Cardiovasc Interv. 2024;103(4):637–649. doi:10.1002/CCD.30970

21. Vogel K, Rojas CN, Greenberg PB, Margo CE, French DD. Impact of the COVID-19 Pandemic on Cataract Surgeries in the United States. Clin Ophthalmol. 2022;16:1601. doi:10.2147/OPTH.S367608

22. N C JR V BE K, et al. Prevalence of cataract and pseudophakia/aphakia among adults in the United States. Arch Ophthalmol. 2004;122(4):487–494. doi:10.1001/ARCHOPHT.122.4.487

